# Sexual dimorphism during LPS-induced systemic inflammation

**DOI:** 10.1101/2025.02.04.25321661

**Authors:** Aron Jansen, Niklas Bruse, Nicole Waalders, Dirk van Lier, Guus Leijte, Jelle Gerretsen, Bas Adriaansen, Teun van Herwaarden, Peter Pickkers, Matthijs Kox

**Affiliations:** Department of Intensive Care Medicine, Radboud university medical center, Nijmegen, The Netherlands; Department of Laboratory Medicine, Radboud university medical center, Nijmegen, The Netherlands

## Abstract

Incidence and outcomes of systemic inflammatory diseases, such as sepsis, differ between men and women, but the underlying mechanisms remain unclear. We examined systemic inflammatory responses in 56 male and 54 female healthy volunteers challenged with 1 ng/kg bacterial lipopolysaccharide (LPS) twice, with an interval of one week. Sex hormones, inflammatory markers, and monocyte RNA expression were determined. Women exhibited higher levels of several proinflammatory mediators than men during the first LPS challenge (TNF +45%, p=0.002; IL-6 +43%, p=0.0009; IP-10 +58%, p<0.0001; G-CSF +60%, p=0.02). Among women, use of hormonal contraceptives was associated with a more pronounced pro-inflammatory response (TNF +60%, p=0.001; IL-8 +30%, p=0.008; IP-10 +37%, p<0.0001). Endogenous concentrations of estrogens and testosterone inversely correlated with the extent of cytokine responses in women, while estrone positively correlated with these responses among men. The magnitude of endotoxin tolerance, exemplified by a significantly blunted cytokine response upon the second LPS challenge, was similar between the sexes, as were monocyte RNA expression patterns. In conclusion, our data indicate that there is considerable sexual dimorphism in systemic inflammation and implicate an important role for sex hormones in regulating immunity.

## Introduction

Sepsis, defined as a life-threatening organ dysfunction caused by a dysregulated host response to infection ^1^, represents a global health care burden to hospitals and to intensive care units (ICUs) in particular. Despite significant advances in supportive care, sepsis mortality rates remain alarmingly high and its incidence continues to rise^2–5^. In fact, sepsis was recently identified as the leading cause of death worldwide, with an estimated 11 million fatalities annually^6^. Unfortunately, over the last decades a plethora of negative sepsis studies have failed to yield effective therapies, and as of today supportive care and source control remain the cornerstone of treatment^7–23^.

One of the primary challenges in managing sepsis is its remarkable heterogeneity, with diverse pathophysiological manifestations across patients. These can include both hyperinflammation, characterized by an overwhelming release of proinflammatory mediators, hemodynamic shock and organ failure as well as immune suppression, marked by excessive anti-inflammatory responses that compromise immune function and increase susceptibility to secondary infections^24,25^. In this light, a solid understanding of the factors that drive these differential disease manifestations is of paramount importance for the development of personalized treatment approaches.

One such factor may be biological sex. Demographic data consistently show that the incidence and outcomes of sepsis differ considerably between men and women, with women generally experiencing more favorable outcomes, including lower ICU admission rates and mortality^26–29^. These sex-specific differences in outcomes are not unique to sepsis, but also hold true for other infectious and systemic inflammatory conditions, such as COVID-19, which affects men more severely than women^30–32^. In contrast, auto-immune diseases such as multiple sclerosis and rheumatoid arthritis are more prevalent among women^33^. These observations suggest that sex-related differences in immune responses may play a significant role, yet robust *in vivo* human data are limited. In this study, we investigate how biological sex influences the immune response in a large cohort of healthy volunteers undergoing repeated experimental endotoxemia, a model that mimics many aspects of both the hyperinflammatory and immunoparalytic phenotypes of sepsis^34,35^.

## Results

### Safety and subject characteristics

All subjects (female n= 54, male n=56) received LPS twice, with an interval of two weeks **(Figure 1A).** All procedures and symptoms associated with the experimental endotoxemia protocol were well tolerated by all subjects, and no (serious) adverse events occurred during the study. To explore potential effects of sex hormones on the response to LPS, female participants were subdivided into two groups based on the use of hormonal contraceptives (HC). Only women who used hormonal contraceptives with a systemic route of administration, such as oral, intramuscular, subcutaneous or transcutaneous contraceptives, were assigned to the HC+ group, whereas intrauterine devices (IUDs) were considered non-systemic contraceptives. Twenty-four female participants were assigned to the HC+ group, leaving 30 women in the HC-group. Baseline demographic characteristics of the study cohort are displayed in **Figure 1B** and reveal no relevant differences between groups.

**Figure 1.**
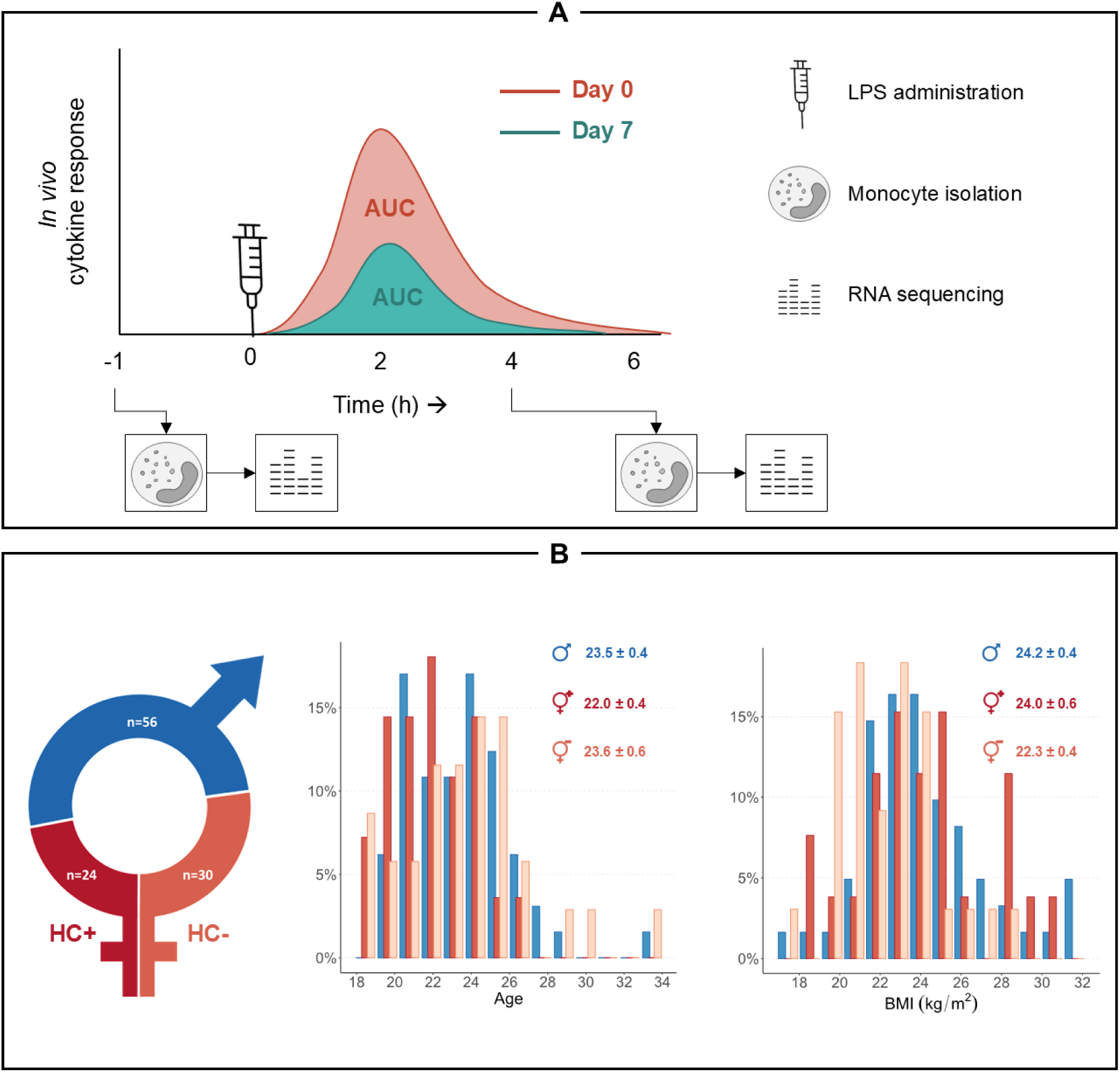
**A:** Schematic representation of the study protocol. **B:** Demographic characteristics of the study cohort. Data are represented as mean ± SEM. HC = hormonal contraceptives.

### Plasma cytokines

The first LPS challenge resulted in a profound increase in all measured inflammatory cytokines in all subjects (**Figure 2** and **Supplementary Figure 1**). Compared to men, women (HC+ and HC-combined) exhibited markedly higher plasma concentrations of proinflammatory cytokines tumor necrosis factor (TNF; 45% higher geometric mean area under curve [AUC] values, p=0.002), interleukin (IL)-6 (+43%, p=0.009), interferon gamma induced protein (IP)-10 (+58%, p<0.0001), and granulocyte colony stimulating factor (G-CSF; +60%, p=0.02), whereas macrophage inflammatory protein (MIP)-1α and monocyte chemoattractant protein (MCP)-1 were not different (+14%, p=0.12 and -4%, p=0.38, respectively). Concentrations of the archetypal anti-inflammatory cytokine IL-10 (+0%, p=0.99) were similar between sexes, whereas the increase in IL-1 receptor antagonist (Ra), another anti-inflammatory cytokine, was more pronounced in women (+95%, p<0.0001).

**Figure 2.**
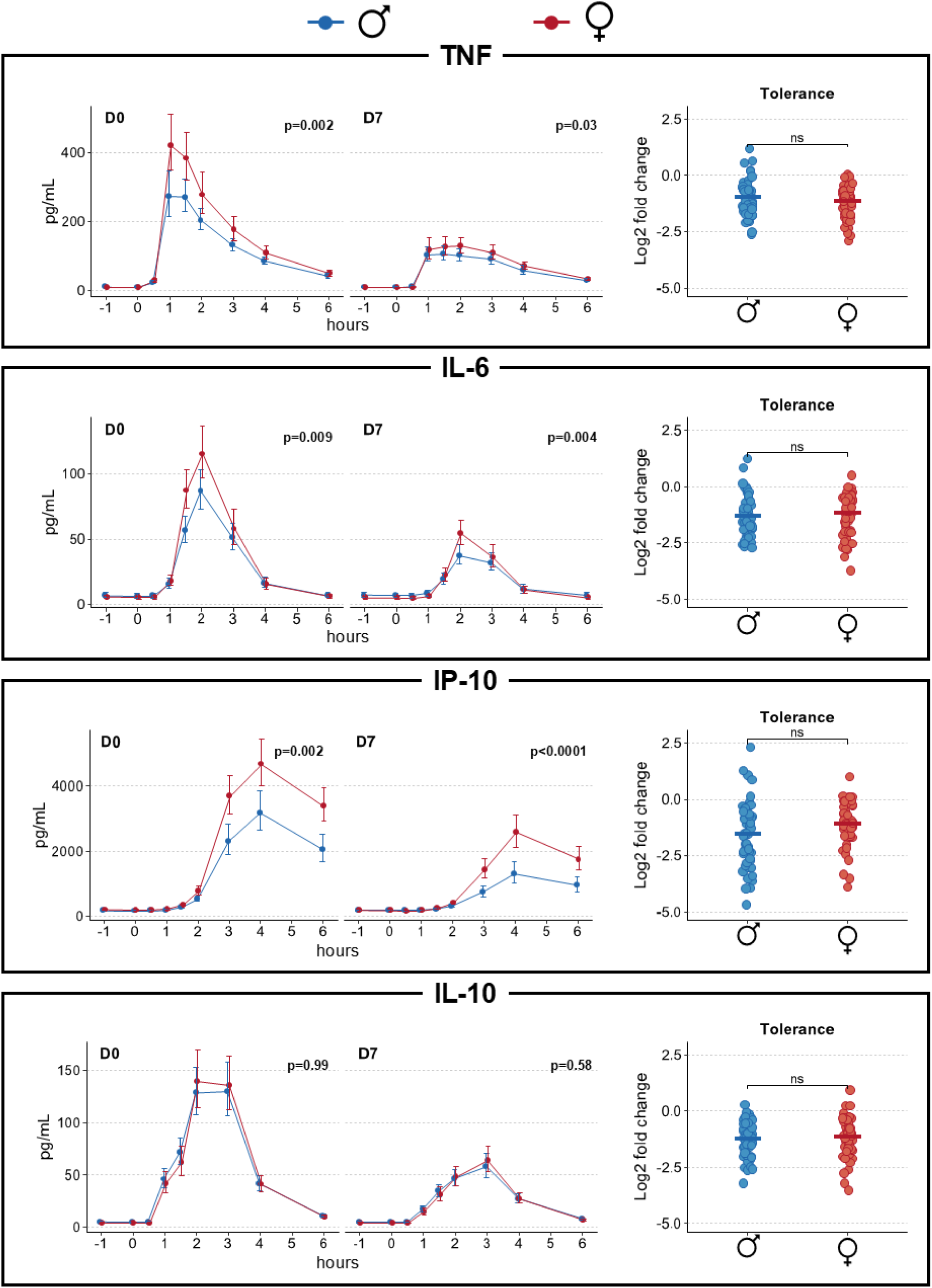
Cytokine time-concentration during the first (D0) and second (D7) LPS challenge, and magnitude of endotoxin tolerance (expressed as the log2 fold change in area under the curve between the two LPS challenge days). Cytokine concentrations are displayed as geometric mean with 95% confidence intervals, whereas tolerance is depicted as jitter plots with the crossbar indicating the mean.

These differences in cytokine responses were also present upon the second challenge, for instance reflected by more pronounced increases in TNF (+25% p=0.03), IL-6 (+51% p=0.004) and IP-10 (+112% p<0.0001) in women compared with men, whereas IL-10 again showed no differences (**Figure 2** and **Supplementary Figure 1**). Furthermore, in both men and women, cytokine responses upon the second challenge were severely blunted as compared to the first challenge (p<0.0001 for all cytokines **Figure 2** and **Supplementary Figure 1**). This phenomenon, known as endotoxin tolerance, was similar in magnitude between sexes for all cytokines **(Figure 2** and **Supplementary Figure 1)**.

### Clinical and hematological parameters

Both LPS challenges led to a transient decrease in blood pressure and an increase in heart rate, body temperature and symptom scores (**Supplementary Figure 2**). The LPS-induced maximum decrease in mean arterial pressure (MAP) was more pronounced in women during both challenges (both p<0.0001, **Supplementary Figure 2**), whereas the compensatory increase in heart rate did not differ between sexes during both challenges (p=0.98 and p=0.22, respectively, **Supplementary Figure 2**). Although the fever response was less pronounced in women than in men upon the first challenge, no differences were observed upon the second LPS challenge (p<0.001 and p=0.11, respectively, **Supplementary Figure 2**). Men and women displayed similar symptom scores over time upon the first challenge (p=0.23), although women exhibited higher scores during the second challenge (p<0.0001, **Supplementary Figure 2**). Moreover, peak scores of several individual symptoms such as headache, nausea and general malaise were higher in women during the first LPS challenge, whereas shivering and general malaise were also more pronounced in women upon the second LPS challenge (**Supplementary Figure 3**). Baseline hemoglobin concentrations and platelet numbers differed significantly between men and women; however, the LPS-induced changes in these parameters over time were similar during both challenges (**Supplementary Figure 4**). In contrast, circulating leukocyte numbers decreased more strongly in women (p<0.0001 for both challenges, **Supplementary Figure 4**), primarily due to a greater reduction in neutrophil counts (p<0.0001 and p=0.0001 during the first and second LPS challenge, respectively, **Supplementary Figure 4**). While the decrease in circulating monocyte counts following LPS administration was comparable between sexes, the monocyte repopulation was notably delayed in women during both challenges (p<0.0001 and p=0.005, respectively, **Supplementary Figure 4**).

### Influence of hormonal contraceptives

Although women in the HC-group still exhibited significantly higher concentrations of several cytokines than men (**Figure 3A** and **Supplementary Figure 5**), the cytokine responses were particularly more pronounced among women of the HC+ group. During both the first and second LPS challenge, use of HC was associated with a more pronounced increase of proinflammatory cytokines TNF (+60%, p=0.001 and +49%, p=0.04, respectively) and IP-10 (+37%, p<0.0001 and +37%, p=0.04, respectively) compared to women in the HC-group (**Figure 3A** and **Supplementary Figure 5)**. Although differences in the IL-10 response did not reach statistical significance upon the first challenge (+41%, p=0.07), HC use was associated with significantly higher concentrations of IL-10 upon the second challenge (+48%, p<0.0001, **Figure 3A** and **Supplementary Figure 5**). Moreover, increases of IL-8, IL-1Ra and G-CSF were also more pronounced in the HC+ group (**Figure 3A** and **Supplementary Figure 5**). No relationship between HC use and the magnitude of endotoxin tolerance was observed (**Figure 3A** and **Supplementary Figure 5).**

**Figure 3.**
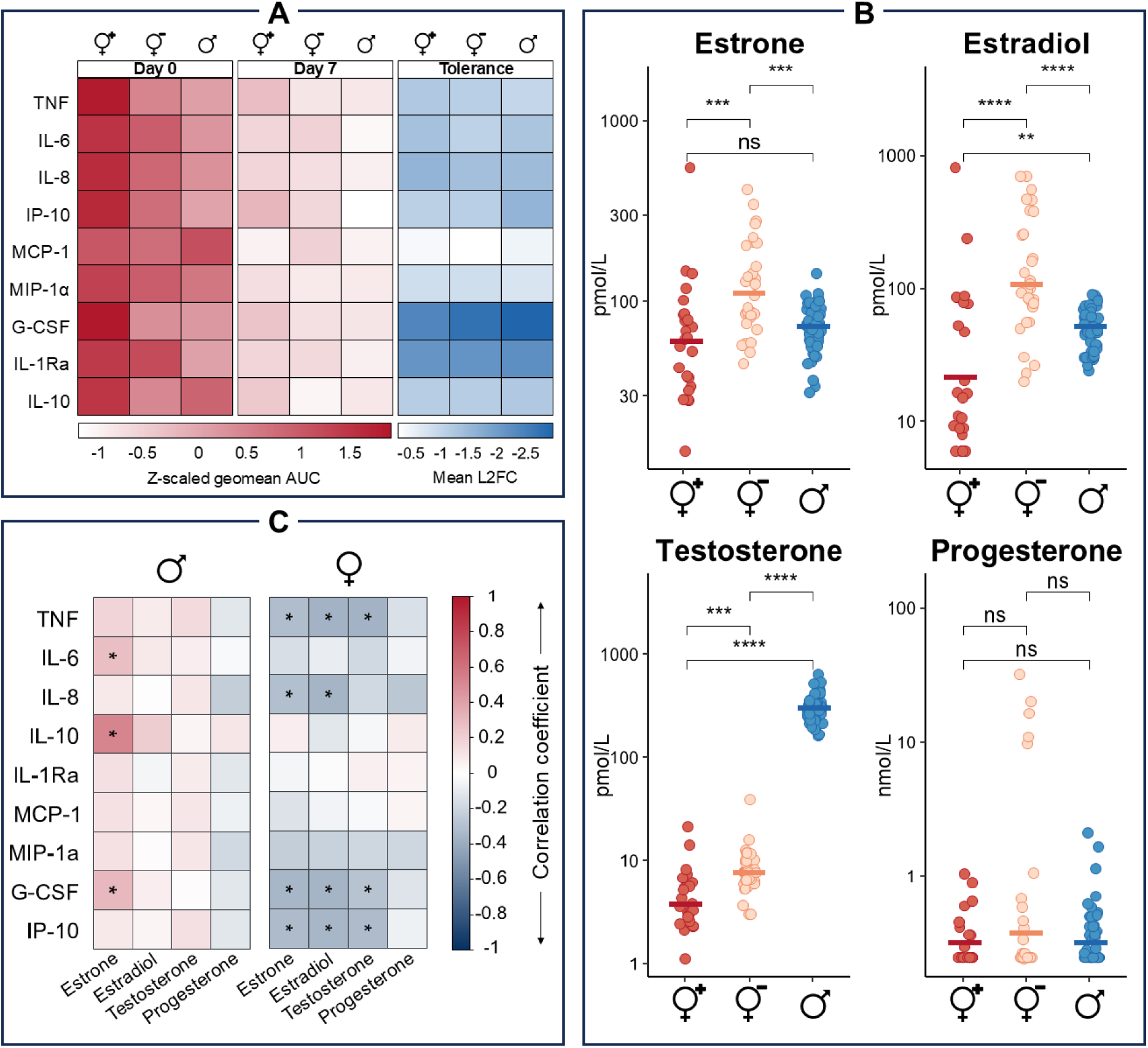
**A:** Heatmap depicting cytokine responses during the two LPS challenges (day 0 and day 7) and magnitude of endotoxin tolerance. Cytokine responses are displayed as Z-scaled geometric mean area under the curve and endotoxin tolerance is depicted as mean log2 fold change between the two LPS challenges. **B:** Differences in baseline concentrations of sex hormones. ♀^+^ = HC+ women, ♀^-^ = HC-women, ♂ = men; ns = not significant; ** = p<0.01; *** = p<0.001; **** = p<0.0001. **C:** Pearson correlations between sex hormone concentrations and area under the curve cytokine responses upon the first LPS challenge (day 0). Correlation coefficients are indicated by color and significant correlations are marked with an asterisk.

Although HC use was not related to differences in MAP and heart rate trajectories over time upon the first challenge (p=0.39 and p=0.69 compared to the HC-group, respectively, **Supplementary Figure 6**), the decrease in MAP was more pronounced in the HC+ group upon the second challenge, whereas the compensatory increase in heart rate was less apparent (both p<0.0001, **Supplementary Figure 6**). During both LPS challenges, the fever response was substantially less pronounced in the HC+ group compared to the HC-group (p=0.0006 and p<0.0001, respectively, **Supplementary Figure 6**). Although the between-group difference in composite symptom score progression over time did not reach statistical significance during both challenges (p=0.053 and p=0.055, respectively, **Supplementary Figure 6**), women in the HC+ group did report higher peak scores of individual symptoms such as general malaise and headache upon the first LPS challenge. However, no such differences between the HC+ and HC-group were observed upon the second challenge (**Supplementary Figure 7**).

### Role of sex hormones

As expected, sex hormone concentrations determined at baseline differed substantially between men and women. For instance, compared to men, women of the HC-group exhibited significantly higher concentrations of estradiol and estrone (p<0.0001 and p=0.0001, respectively, **Figure 3B**), whereas testosterone concentrations were lower (p<0.0001, **Figure 3B**). Differences in progesterone did not reach statistical significance (p=0.06, **Figure 3B**). Use of HC was associated with suppression of endogenous estrogens, resulting in lower concentrations of estradiol and estrone (p<0.0001 and p=0.001 compared to the HC-group, respectively, **Figure 3B**). Testosterone concentrations were also lower in the HC+ group (p=0.0002, **Figure 3B**) than in the HC-group, and a trend towards lower progesterone concentrations was observed (p=0.06, **Figure 3B**).

Correlations between sex hormone concentrations and cytokine responses during the first LPS challenge were determined among men and women separately to preclude group-based effects. Among men, estrone concentrations positively correlated with IL-6, IL-10 and G-CSF responses, although no significant relationships between estradiol and any of the cytokines were present (**Figure 3C**). In women, concentrations of the estrogens estrone and estradiol correlated inversely with TNF, IL-8, G-CSF, and IP-10 responses (**Figure 3C** and **Supplementary Figure 8**). No associations between testosterone concentrations and cytokine responses were found among men (**Figure 3C** and **Supplementary Figure 8**). In contrast, testosterone concentrations were inversely correlated with several pro-inflammatory cytokines in women (TNF, G-CSF, and IP-10, **Figure 3C** and **Supplementary Figure 8**). No relationships between progesterone concentrations and cytokine responses were found among both men and women.

### Monocyte RNA expression

To investigate sex-specific differences in RNA expression, monocytes were isolated at baseline (T0) and four hours following the first LPS administration (T4), and RNA sequencing was performed (**Figure 1A**). After filtering genes with low counts across all samples, 14,564 genes remained for downstream analysis. An exploratory principal component analysis revealed that the first component (PC1) was mainly dependent on the timepoint (and thus driven by LPS administration), accounting for 39% of the observed variance in this dataset. PC2 accounted for 14% of the observed variance and was mainly driven by sex (**Figure 4A**). Differential expression analysis between samples obtained at T0 and T4 revealed that LPS administration resulted in more than 11,000 differentially expressed genes across men and women (**Figure 4B**). With an overlap of 90.5% in upregulated genes and 88.4% in downregulated genes, the monocyte transcriptomic response to LPS administration proved highly similar between the two sexes (**Figures 4B** and **4C**). Next, the 20 most upregulated and downregulated genes were selected based on their log2 fold change and their expression levels on T4 were plotted in a heatmap, again displaying great similarity between men and women (**Figure 4D**). Finally, genes which were differentially expressed between men and women on T4 were explored using Reactome pathway analysis (**Figure 4E**). Several gene sets associated with hemostasis, signal transduction, developmental biology, reproduction, chromatin organization, and DNA localization were significantly more enriched in female subjects, whereas gene sets related to protein localization were less enriched in women. However, no significant differences in pathways associated with the immune system and cellular responses to stimuli were observed.

**Figure 4.**
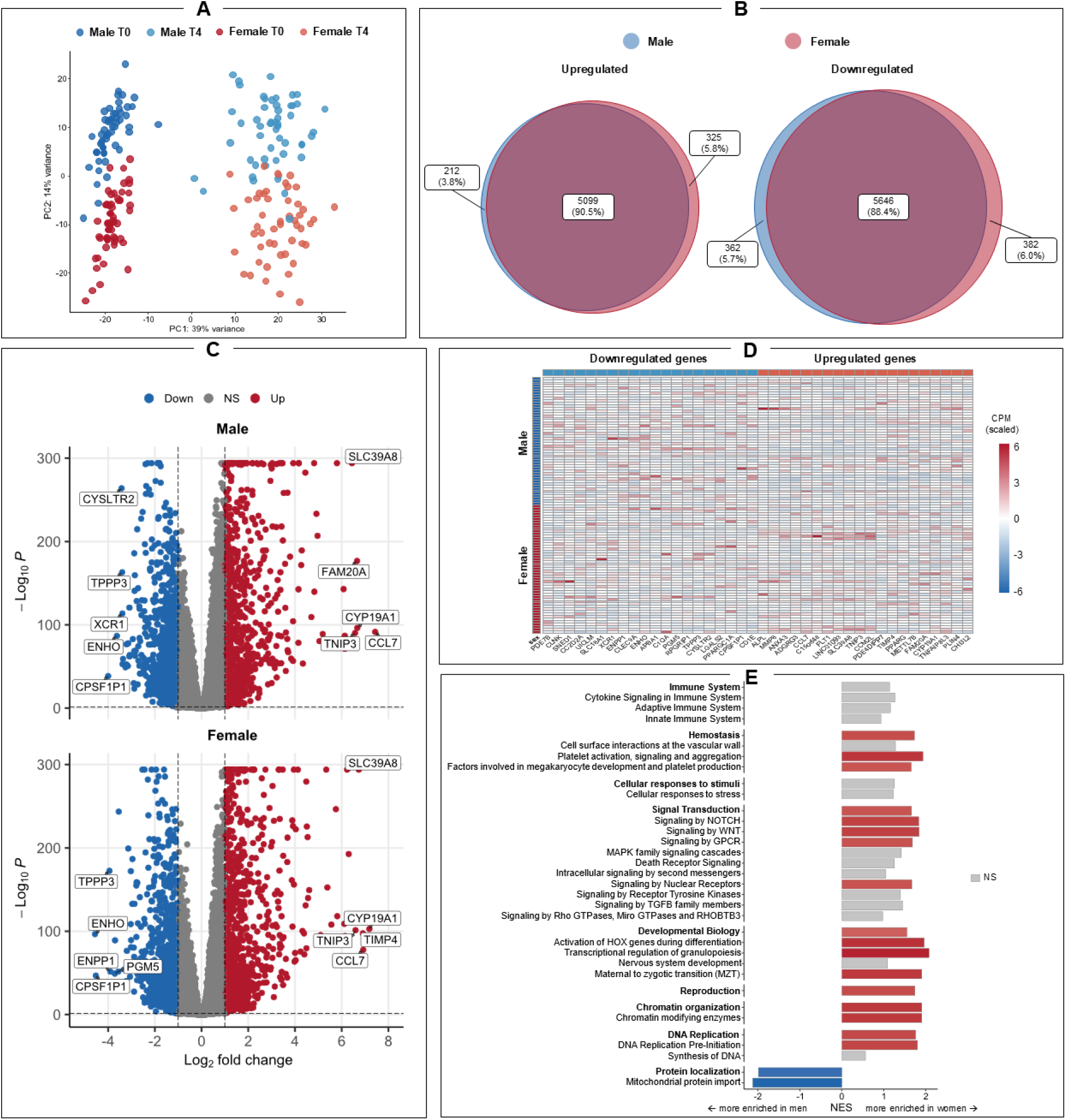
**A:** Principal component analysis of the variance in gene expression in monocytes isolated at baseline (T0) and four hours (T4) after the first LPS challenge (day 0). **B:** Venn-diagrams displaying the overlap in significantly upregulated and downregulated genes between male and female subjects. **C:** Volcano-plots of differentially expressed genes between monocytes isolated at T0 and T4 for male and female participants. **D:** Heatmap depicting normalized counts per million (CPM) at T4 of the top 20 upregulated and downregulated genes in all subjects of the study cohort. **E:** Bar-plot displaying differences in gene-set enrichment between male and female subjects (Reactome analysis). Nested enrichment scores (NES) are indicated by color, and non-significantly enriched terms are displayed in grey.

## Discussion

Sepsis and numerous other inflammatory diseases exhibit distinct incidence rates and outcome statistics among men and women, suggesting the presence of sexual dimorphism in immunity. Although the origins of these differences have been extensively investigated in preclinical (animal) models and *in vitro* studies, comprehensive *in vivo* data from human subjects remain limited. In the present work, we demonstrate that women mount a more pronounced proinflammatory response to intravenous administration of bacterial LPS than men. Furthermore, our findings indicate that hormonal contraceptive use is strongly associated with the magnitude of the immune response, highlighting a significant influence of sex hormones in regulation of the immune response. Finally, our study signifies that despite these marked sex-based differences in the magnitude of the LPS-induced immune response, monocyte RNA expression patterns were largely similar between male and female participants.

Our results are in agreement with previous endotoxemia studies that also reported a more pronounced proinflammatory response in women^29,36–38^. However, neither the role of sex hormones and hormonal contraceptives, nor transcriptomic responses had been studied to date. The use of hormonal contraceptives was associated with more pronounced cytokine responses. In line, estrone concentrations were correlated with stronger cytokine responses in men. In contrast, estrogens were inversely related to cytokine responses among women. Perhaps counterintuitively, this may actually support the pro-inflammatory effects of estrogens found in men, as hormonal contraceptives used by a large proportion of women in our cohort strongly suppress endogenous estrogens, which our data clearly show as well. The effects of estrogens on immune function are not uncontested in literature^39^ and appear to be concentration-dependent: low or physiological concentrations (such as those evoked by hormonal contraceptive use) tend to promote inflammation in animal and *in vitro* studies, while higher concentrations (such as observed during pregnancy) are generally thought to be anti-inflammatory^39,40^. The putative proinflammatory effects of hormonal contraceptives are further supported by clinical studies reporting elevated levels of C-reactive protein and other acute phase proteins in young women using oral contraceptives^41,42^. Additionally, the use of hormonal contraceptives has been linked to an increased susceptibility to several autoimmune diseases, including multiple sclerosis, Crohn’s disease, and lupus^43^. Based on these effects, one might hypothesize that hormonal contraceptives could have therapeutic potential in managing sepsis-induced immune suppression. However, since our study found no impact of hormonal contraceptives on the development of endotoxin tolerance, our results do not directly support this hypothesis. Testosterone concentrations were inversely correlated with several proinflammatory cytokines in female participants, supporting a broader body of literature that suggests an anti-inflammatory role of androgens^39^. Although no such correlations were observed in male subjects, this may be attributed to a threshold effect, given that our study comprised a highly homogeneous group of young, healthy volunteers with high testosterone levels. This may have obscured evidencing relationships between testosterone and immune function among men. This hypothesis is consistent with findings from a study in male COVID-19 patients, where inverse correlations between testosterone concentrations and several proinflammatory cytokines were found^44^.

The transcriptomic responses of monocytes upon an LPS challenge were largely similar between men and women, only differing in magnitude rather than specific molecular pathways. These results align with a clinical cohort study of sepsis patients, which also reported an 80% overlap in leukocyte gene expression between male and female patients^45^. It appears plausible that the apparent lack of a genomic substrate for the observed differences in cytokine production reflects the highly conserved nature of host defense pathways.

Our study has several strengths. Unlike immunological studies in sepsis patients, which are often confounded by factors such as age, medical history, medication use, disease severity and source of infection, the experimental endotoxemia model offers a highly standardized and reproducible framework to investigate systemic inflammation in humans *in vivo*. Furthermore, while experimental endotoxemia studies are often constrained by small sample sizes due to their cost and labor-intensive nature, we included over 100 participants, enabling a more comprehensive exploration of the factors underlying immunological differences between male and female participants. However, although the homogeneity of our study population is a benefit in terms of confounding factors, it clearly also represents a limitation, as results from our study may not be generalizable to populations with broader demographic diversity. Moreover, the inflammatory response induced by experimental endotoxemia is relatively acute and transient, which may limit its translatability to diseases and conditions characterized by sustained systemic inflammation. Nevertheless, previous work has shown that many hallmarks of the immune response in sepsis, including hyperinflammatory and immunosuppressive mechanisms, are also present in the endotoxemia model^35^. Finally, RNA sequencing was only performed in monocytes and it cannot be excluded that differential transcriptomic responses between men and women are present in other immune cells, such as lymphocytes, neutrophils, tissue-resident macrophages, or endothelial cells.

In conclusion, we demonstrate that women mount a markedly more pronounced proinflammatory response during experimental endotoxemia than men, that the observed differences in cytokine production do not arise from distinct underlying molecular pathways, and that responses are strongly influenced by sex hormones, supporting an important immunoregulatory role for these mediators.

## Methods

### Subjects

After approval of the local ethics committee (Medical Ethical Committee Oost-Nederland; reference no. NL68166.091.18), 56 male and 54 female healthy volunteers between 18-35 years old were recruited for the endotoxemia study. All subjects provided written informed consent and were included after medical history, physical examination, routine laboratory tests, and a 12-lead electrocardiogram did not reveal any abnormalities. Smoking, use of any medication (with exception of contraceptives), previous participation in experimental human endotoxemia, or signs of acute illness within two weeks prior to the start of the study were exclusion criteria. All study procedures were performed in compliance with the declaration of Helsinki, including the latest revisions.

### Study design

We performed a prospective experimental cohort study, the design of which is depicted in **Figure 1A**. All subjects were intravenously challenged with bacterial lipopolysaccharide (LPS) twice, on study days zero and seven. The first challenge served to induce a systemic inflammatory response and the development of endotoxin tolerance. The second challenge served to quantify the degree of endotoxin tolerance, which is reflected by a blunted cytokine response upon the second challenge as compared to the first challenge^35,46–48^. To gain insight into the effects of contraceptives on the immune response, questionnaires regarding contraceptive methods were sent out to all participants prior to the first LPS challenge. In female participants, pregnancy was excluded prior to LPS administration on both days by using a human chorionic gonadotropin (hCG) urine dipstick test. Other than that, there were no differences in the protocol between female and male participants.

### Experimental endotoxemia

Experimental endotoxemia procedures were performed as described extensively elsewhere ^34^. In short: during both LPS challenge days, subjects were admitted to the research unit of the Intensive Care Department of the Radboud university medical center for 8 hours. Subjects had to refrain from alcohol and caffeine (24 hours), and food and drinks (12 hours) prior to LPS administration. Upon admission, subjects were weighed to determine the total dose of LPS to be administered (1 ng/kg bodyweight). An intravenous cannula was placed in an antebrachial vein to administer fluids and LPS. A radial artery catheter (BD Infusion Therapy Systems, Sandy, USA) was placed under ultrasound guidance to allow serial blood sampling and continuous monitoring and collection of hemodynamic data. Prior to LPS administration, prehydration fluids (1.5 liters of 2.5% glucose / 0.45% sodium chloride in 45 minutes) were administered to reduce the risk of vasovagal responses ^49^. Thereafter, a bolus of 1 ng/kg LPS (*E. coli Type O113*, Lot no. 94332B1; List Biological Laboratories, Campbell, USA) was administered. Hydration fluids (2.5% glucose / 0.45% sodium chloride) were administered at a rate of 150 mL/h continuously throughout the experiment.

### Clinical parameters

During hospitalization, heart rate and intra-arterial blood pressure were continuously monitored using a 4-lead electrocardiogram (M50 Monitor, Philips, Eindhoven, the Netherlands), and a radial artery pressure transducer (Edward Lifesciences, Irvine, USA) connected to the Philips monitor, respectively. Hemodynamic data were sampled every 5 seconds using in-house developed software. Every 30 minutes, core temperature was measured with a tympanic thermometer (FirstTemp Genius 2, Covidien, Dublin, Ireland). Flu-like symptoms (headache, nausea, cold shivers, muscle and back pain) were scored using a numeric six-point Likert scale (0 = no symptoms, 5 = worst ever experienced) whereas general malaise was scored on an eleven-point Likert scale (0 = no symptoms, 10 = worst ever experienced). A total composite symptom score was then calculated as the sum of all previous elements, resulting in a total symptom score ranging from 0 to 35.

### Sex hormone measurements

Sex hormone profiles were assessed at baseline (before the first LPS challenge). To this end, blood samples were collected in serum clot activator tubes and allowed to clot for 30 minutes. Tubes were subsequently centrifuged (10 min, 2000g, 4°C) and serum was stored at -80°C until analysis. Concentrations of testosterone, progesterone, estrone and estradiol were determined batchwise using liquid chromatography – mass spectrometry (LC-MS).

### Cytokine measurements

For plasma cytokines, EDTA-anticoagulated blood was centrifuged (10 min, 2000g, 4°C) directly after withdrawal and plasma was stored at -80°C until analysis. Concentrations of tumor necrosis factor (TNF), interleukin (IL)-1 receptor antagonist (Ra), IL-6, IL-8, IL-10, macrophage inflammatory protein (MIP)-1α, monocyte chemoattractant protein (MCP)-1, granulocyte colony stimulating factor (G-CSF) and interferon-γ induced protein (IP)-10 were determined batchwise using a simultaneous Luminex assay (Milliplex, Millipore, Billerica, USA).

### RNA sequencing

Classical monocytes (CD14+CD16-) were isolated at baseline (T0) and four hours after LPS administration (T4). To this end, PBMCs were first isolated as described above. Subsequently, classical monocytes were isolated by immunomagnetic negative selection using a monocyte isolation kit (EasySepTM Human Monocyte Isolation Kit, STEMCELL Technologies, Cologne, Germany) and RNA was isolated using the RNeasy kit (Qiagen, Hilden, Germany) as per the manufacturer’s instructions. Sequencing of the isolated samples was performed by BGI Genomics (Shenzhen, China) using their proprietary DNBseq^TM^ platform. Using a pipeline developed in-house by the Center for Molecular and Biomolecular Informatics (CMBI) of Radboud university medical center, the resulting paired-end reads (.fastQ files) were trimmed using Trim Galore! v0.4.5 to remove low-quality bases and adapters, quality checked using MultiQC and subsequently mapped against the GRCh38 reference transcriptome using STAR v2.6.0a ^50–52^. Gene counts were quantified using HTSeq v0.11.0, resulting in gene counts for 58,740 genes ^53^. Genes with low counts across all samples were filtered using the default ‘filterByExpr()’ function from edgeR v4.0.2 ^54^. Next, differential expression analysis between samples obtained at T0 and T4 was performed using DESeq2 v1.42.0. Finally, to further explore potential between-sex differences in the transcriptomic response to LPS, pathway analysis was performed on differentially expressed genes between male and female subjects on T4 using ReactomePA v1.46.0 ^55^.

### Statistical analysis

Distribution of data was tested for normality using Shapiro-Wilk’s test, and data with a non-Gaussian distribution were log-transformed prior to statistical testing. Data with a Gaussian distribution are presented as mean ± standard error (SEM), and non-parametric data as geometric mean (95%-confidence interval) or median [interquartile range]. The area under the time-concentration curve (AUC) was used as an integral measure of cytokine responses over time during each LPS challenge day. In the text, percentage difference in cytokine AUCs are reported to illustrate differences in responses to LPS. The degree of endotoxin tolerance was calculated per cytokine as the log2 fold change in AUC between the two LPS challenges. Differences in baseline characteristics were analyzed using unpaired student’s t-tests. Repeated measures two-way analysis of variance (ANOVA, time*group interaction term) was used to analyze between-group differences over time within the same LPS challenge day. Within-group differences between the two LPS challenge days were compared using paired student’s t-tests. Pearson’s correlation analysis was used. A two-tailed p-value <0.05 was considered statistically significant, and results were corrected for multiple testing using the false discovery rate according to Benjamini and Hochberg where applicable ^56^. All statistical analyses were performed using GraphPad Prism version 8 (GraphPad Software, San Diego, USA) and R version 4.3.2 (The R Foundation for Statistical Computing, Vienna, Austria).

## Data Availability

All data produced in the present study are available upon reasonable request to the authors

## Supplementary Figures

**Supplementary Figure 1.**
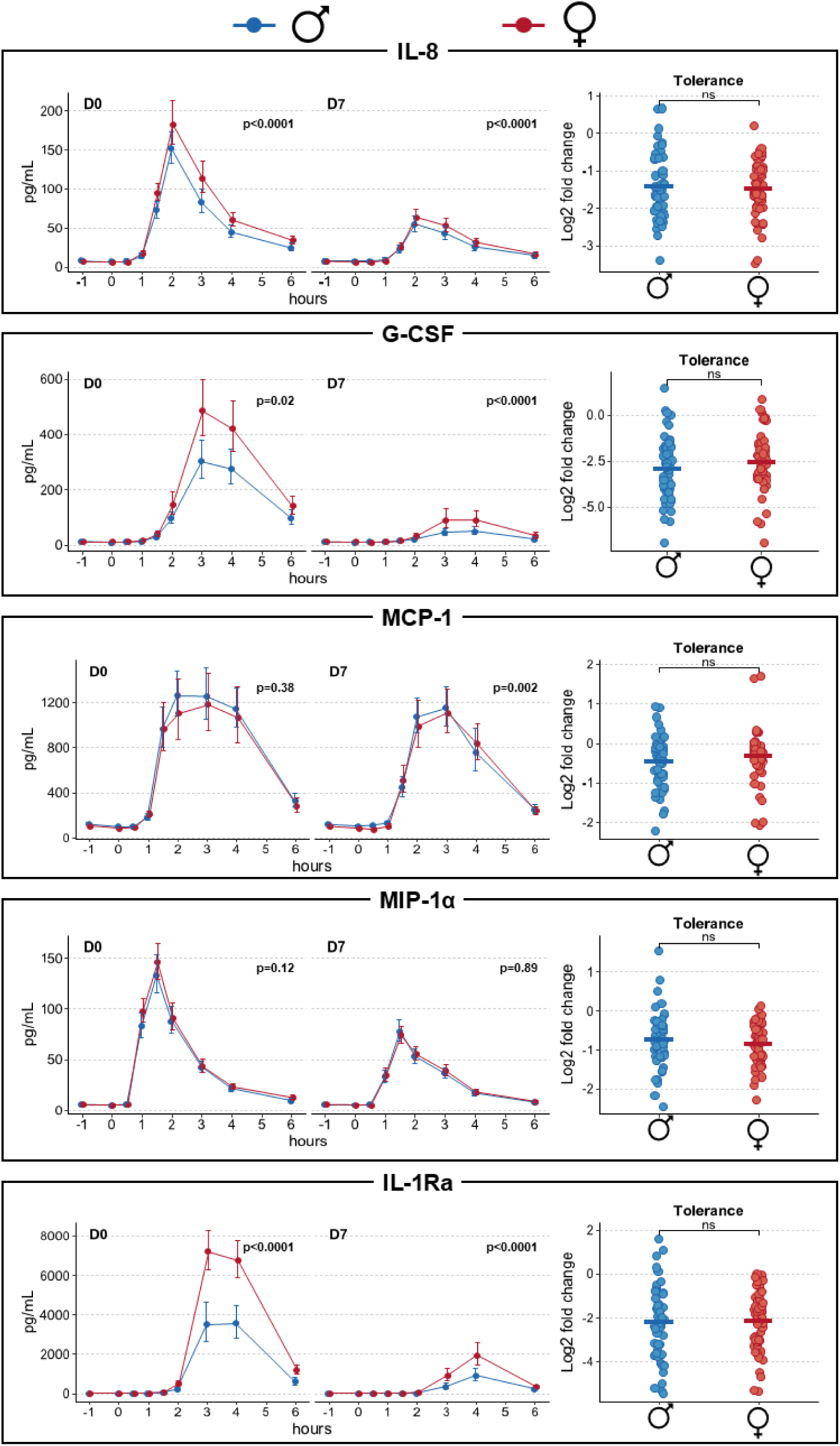
Cytokine time-concentration curves during the first (D0) and second (D7) LPS challenge, and magnitude of endotoxin tolerance (expressed as the log2 fold change in area under the curve between the two LPS challenge days). Cytokine concentrations are displayed as geometric mean with 95% confidence intervals, whereas tolerance is depicted in jitter plots with the crossbar indicating the mean.

**Supplementary Figure 2.**
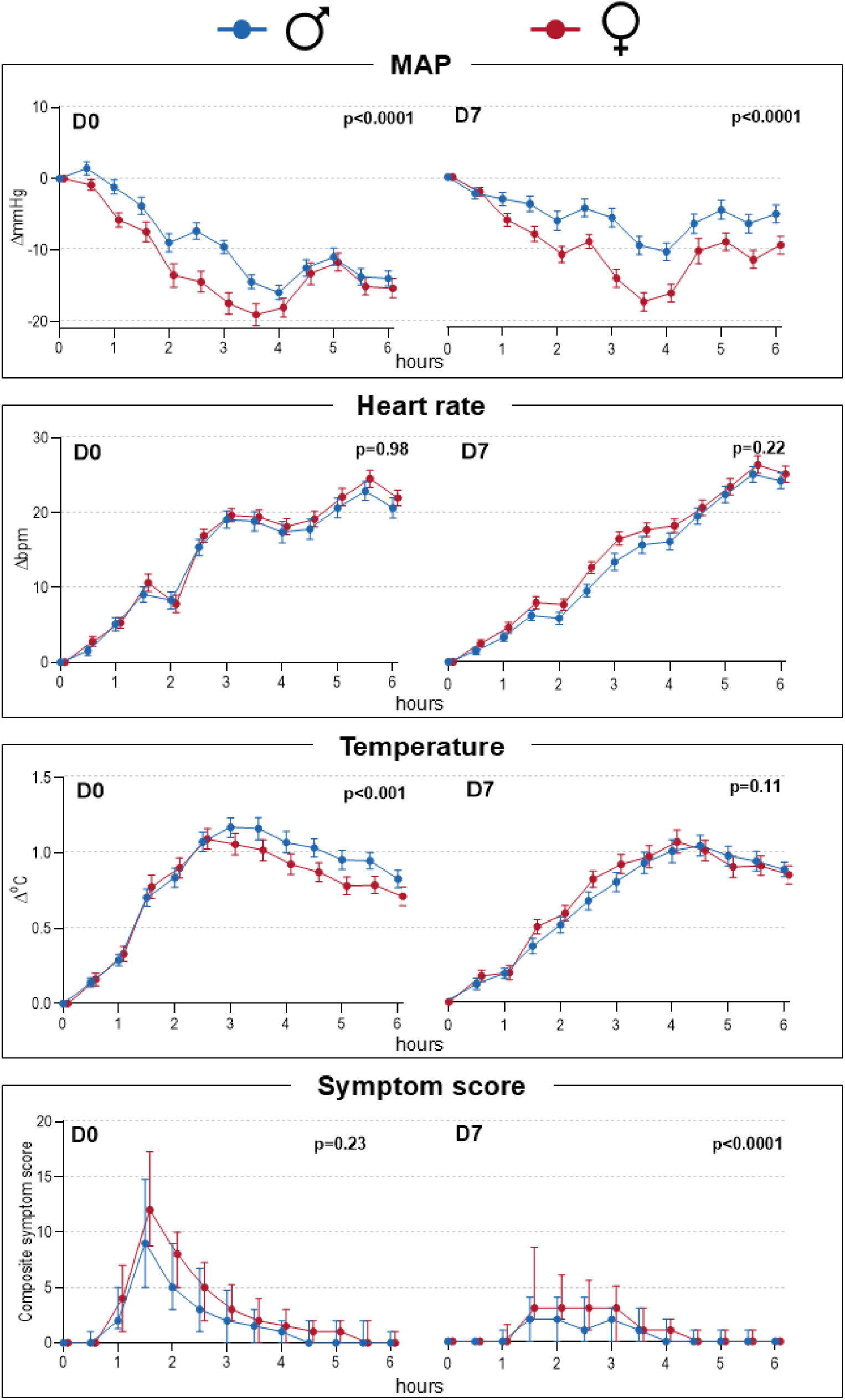
LPS-induced changes over time in blood pressure, heart rate, body temperature and composite symptom scores during the first (D0) and second (D7) LPS challenge. Blood pressure, heart rate and temperature are displayed as mean ± SEM, whereas symptom scores are displayed as median and interquartile range. MAP = mean arterial pressure; mmHg = millimeters of mercury; bpm = beats per minute; °C = degrees Celsius.

**Supplementary Figure 3.**
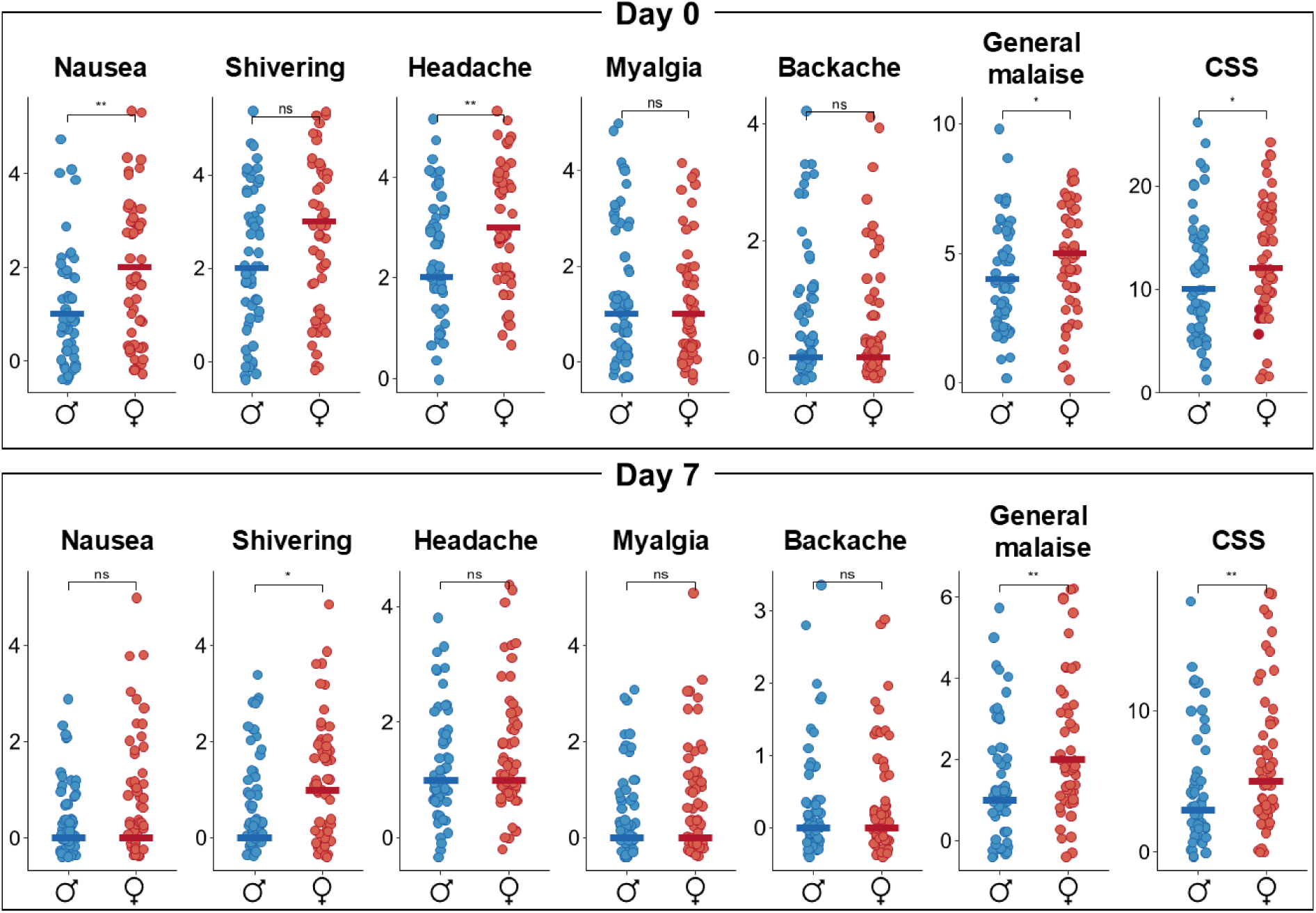
Differences in peak scores of individual symptoms and composite symptom score during the first (day 0) and second (day 7) LPS challenge. Data displayed as jitter plots with the crossbar indicating the median. CSS = composite symptom score. ns = not significant; * = p≤0.05; ** = p<0.01.

**Supplementary Figure 4.**
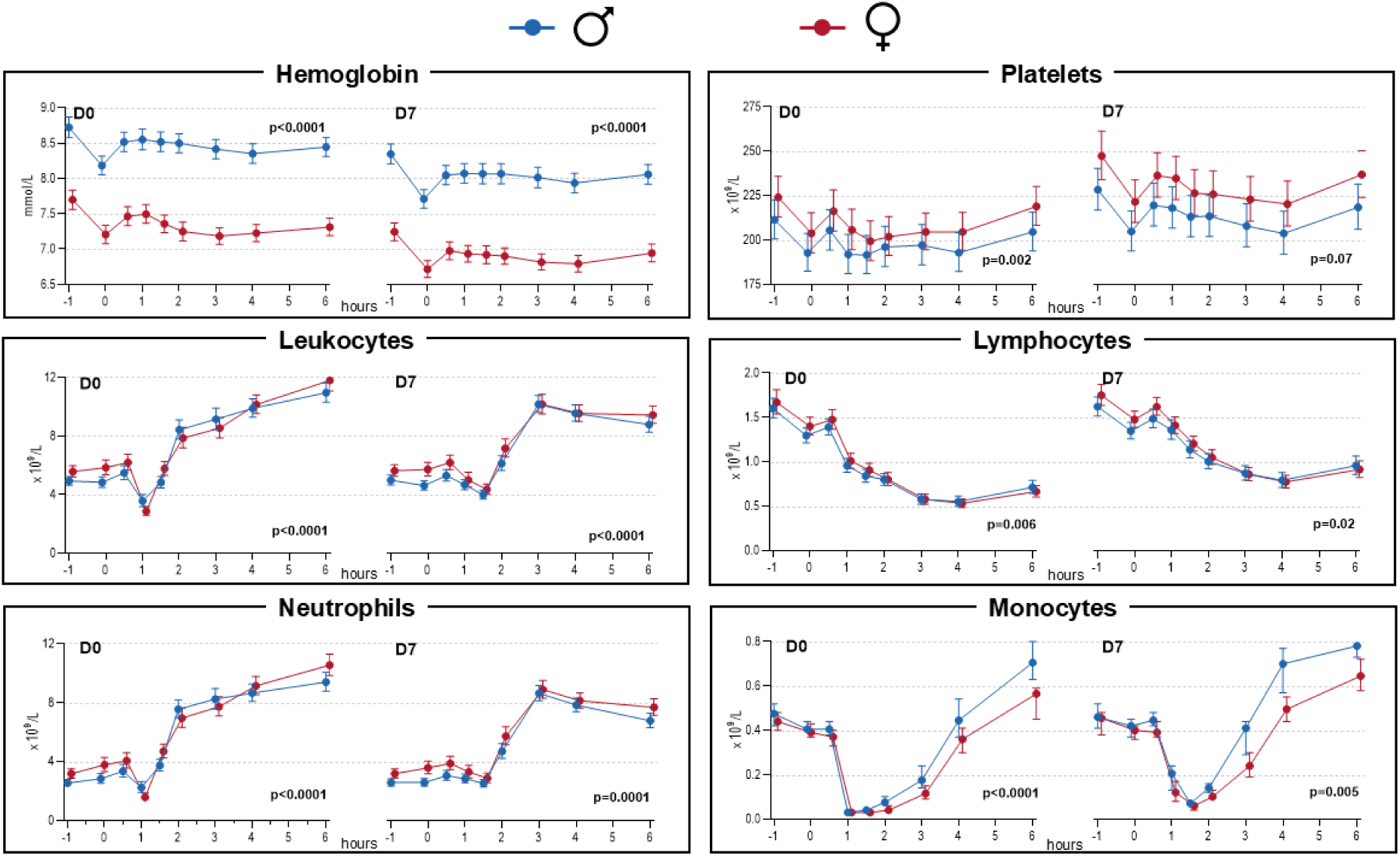
Time-concentration curves of hemocytometry parameters during the first (D0) and second (D7) LPS challenge. Data displayed as mean ± SEM.

**Supplementary Figure 5.**
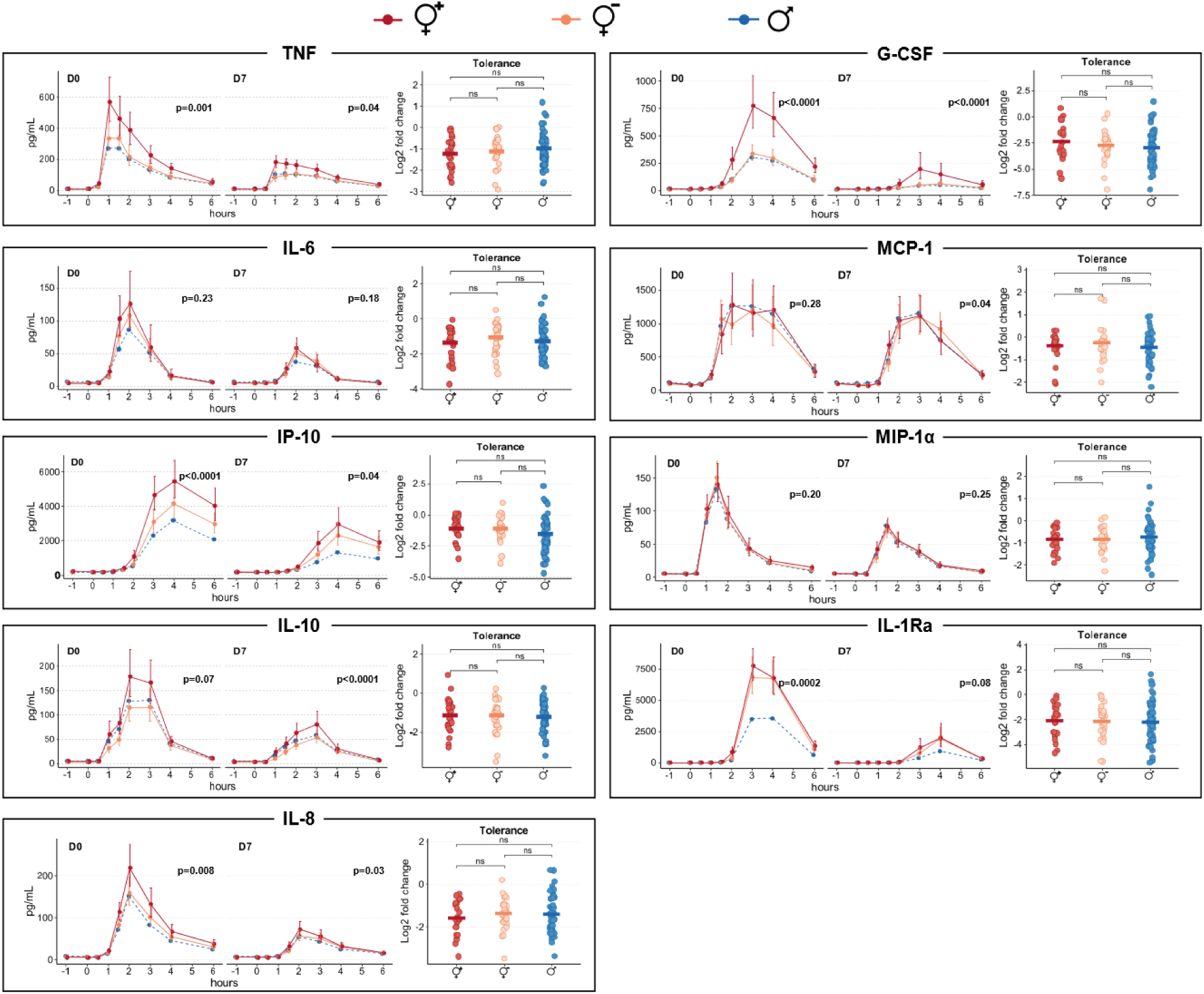
Cytokine time-concentration curves on during the first (D0) and second (D7) LPS challenge, and magnitude of endotoxin tolerance (expressed as the log2 fold change in area under the curve between the two LPS challenge days). Cytokine concentrations are displayed as geometric mean with 95% confidence intervals, whereas tolerance is depicted in jitter plots with the crossbar indicating the mean. ♀^+^ = HC+ women; ♀^-^ = HC-women; ♂ = men; ns = not significant.

**Supplementary Figure 6.**
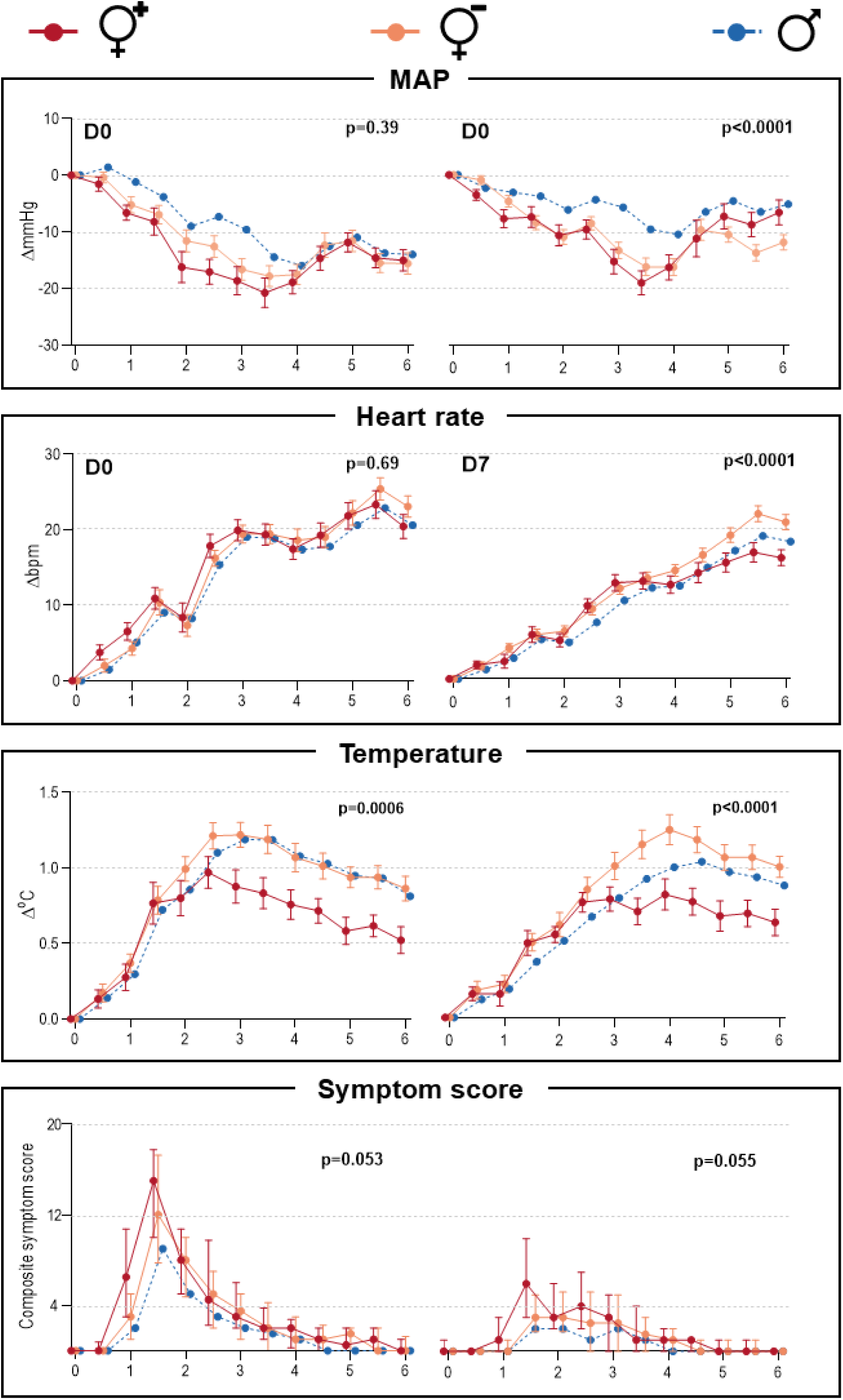
LPS-induced changes over time in blood pressure, heart rate, body temperature and composite symptom scores during the first (D0) and second (D7) LPS challenge. Blood pressure, heart rate and temperature are displayed as mean ± SEM, whereas symptom scores are displayed as median and interquartile range. MAP = mean arterial pressure; mmHg = millimeters of mercury; bpm = beats per minute; °C = degrees Celsius; ♀^+^ = HC+ women; ♀^-^ = HC-women; ♂ = men.

**Supplementary Figure 7.**
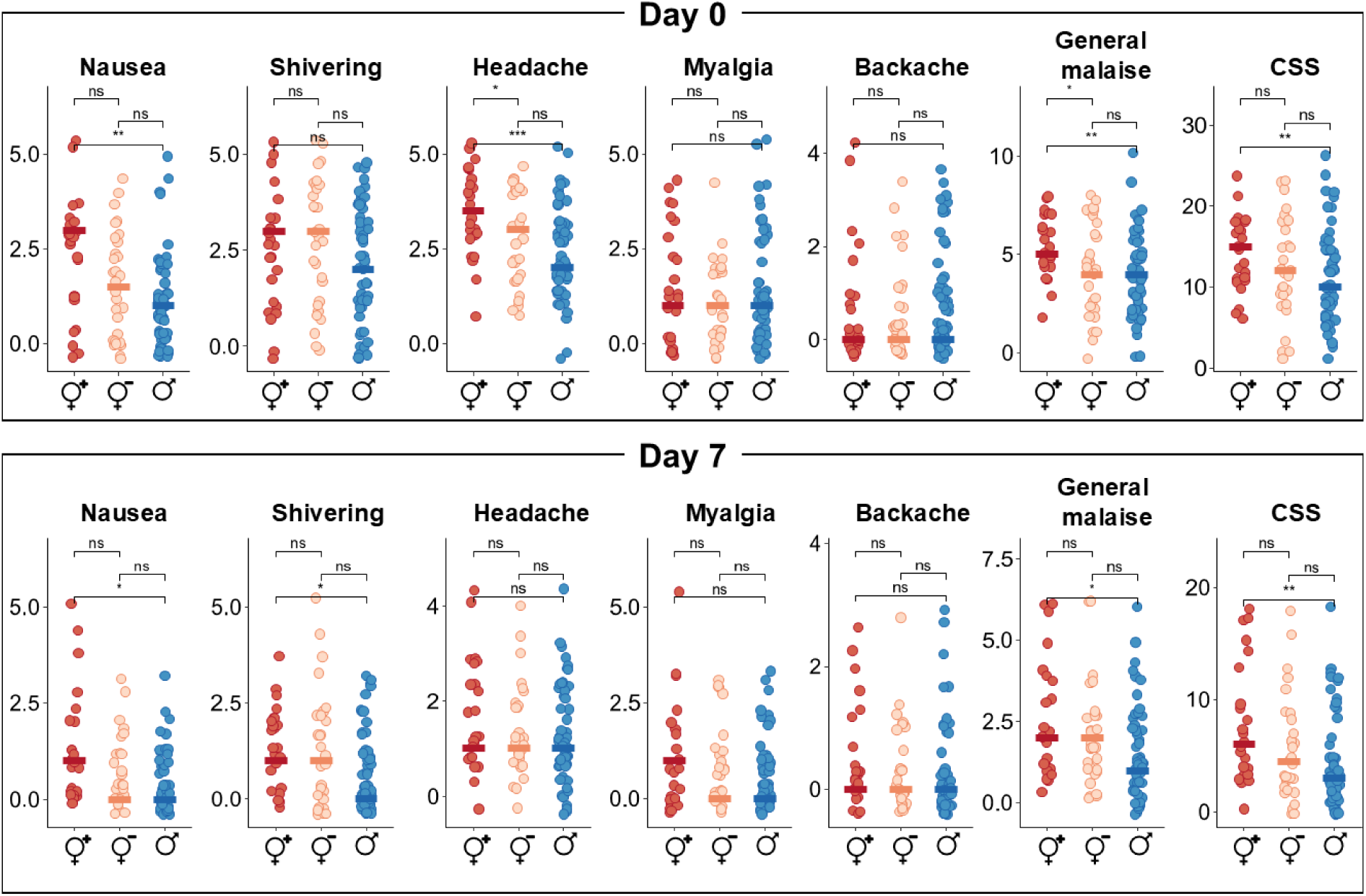
Differences in peak scores of individual symptoms and composite symptom score during the first (day 0) and second (day 7) LPS challenge. Data displayed as jitter plots with the crossbar indicating the median. CSS = composite symptom score. ns = not significant; * = p≤0.05; ** = p<0.01; ♀^+^ = HC+ women; ♀^-^ = HC-women; ♂ = men.

**Supplementary Figure 8.**
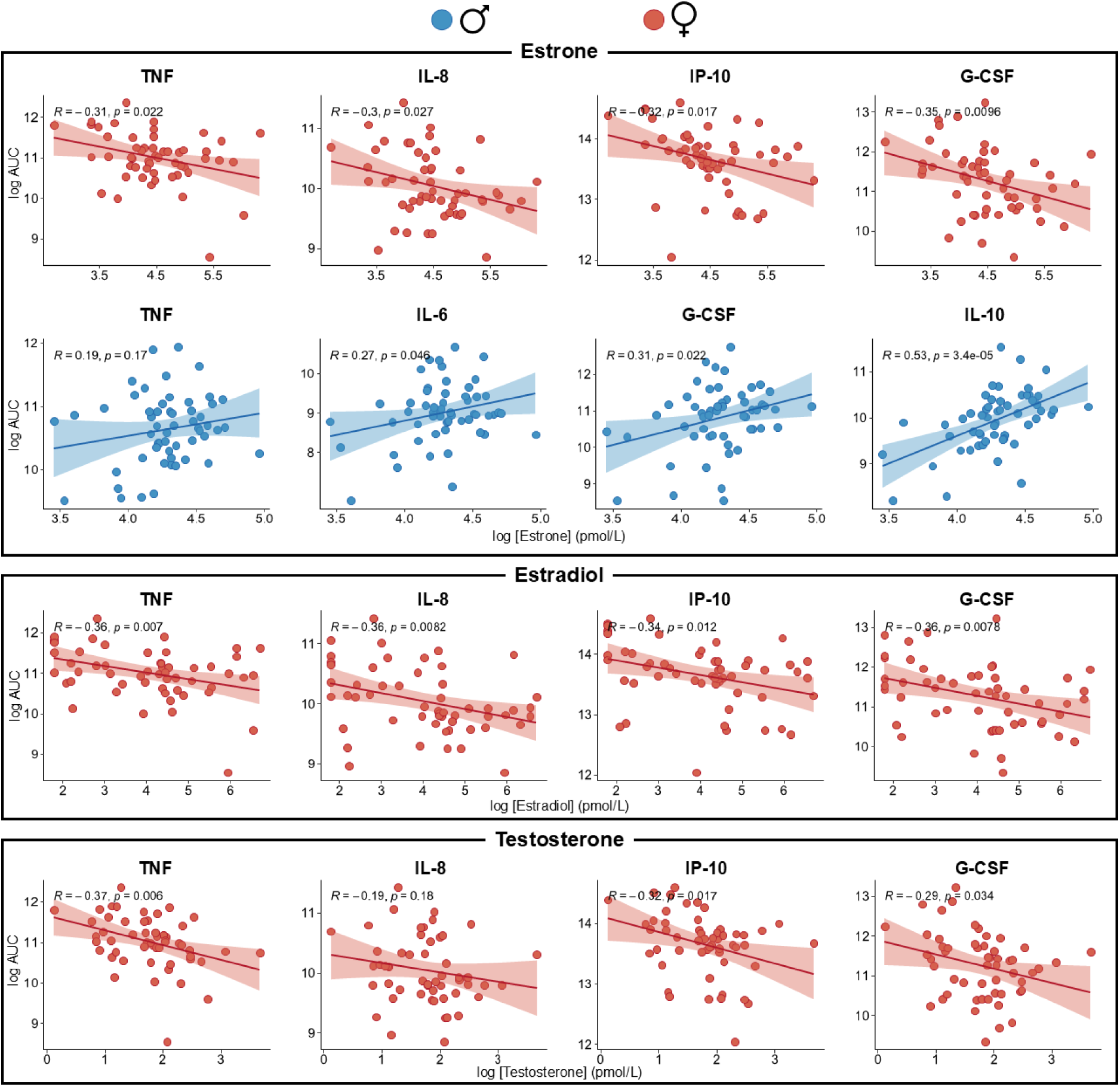
Scatterplots displaying Pearson correlations between baseline sex hormone concentrations and area under the curve (AUC) of inflammatory cytokines during the first LPS challenge.

## Notes

### Competing Interest Statement

The authors have declared no competing interest.

### Clinical Trial

NCT06801873

### Funding Statement

This study did not receive any funding

### Author Declarations

The Medical Ethical Committee Oost-Nederland gave ethical approval for this work (reference no. NL68166.091.18)

